# Comparison of Multimodal Deep Learning Approaches for Predicting Clinical Deterioration in Ward Patients

**DOI:** 10.1101/2025.03.06.25322855

**Authors:** Charles A. Kotula, Jennifer Martin, Kyle A. Carey, Dana P. Edelson, Dmitriy Dligach, Anoop Mayampurath, Majid Afshar, Matthew M. Churpek

## Abstract

**Objective:** Implementing machine learning models to identify clinical deterioration on the wards is associated with improved outcomes. However, these models have high false positive rates and only use structured data. Therefore, we aim to compare models with and without information from clinical notes for predicting deterioration.

**Materials and Methods:** Adults admitted to the wards at the University of Chicago (development cohort) and University of Wisconsin-Madison (external validation cohort) were included. Predictors consisted of structured and unstructured variables extracted from notes as Concept Unique Identifiers (CUIs). We parameterized CUIs in five ways: Standard Tokenization (ST), ICD Rollup using Tokenization (ICDR-T), ICD Rollup using Binary Variables (ICDR-BV), CUIs as SapBERT Embeddings (SE), and CUI Clustering using SapBERT embeddings (CC). Each parameterization method combined with structured data and structured data-only were compared for predicting intensive care unit transfer or death in the next 24 hours using deep recurrent neural networks.

**Results:** The study included 506,076 ward patients, 4.9% of whom experienced the outcome. The SE model achieved the highest AUPRC (0.208), followed by CC (0.199) and the structured-only model (0.199), ICDR-BV (0.194), ICDR-T (0.166), and ST (0.158). The CC and structured-only models achieved the highest AUROC (0.870), followed by ICDR-T (0.867), ICDR-BV (0.866), ST (0.860), and SE (0.859).

**Discussion:** A multimodal model combining structured data with embeddings using SapBERT had the highest AUPRC, but performance was similar between models with and without CUIs.

**Conclusion:** The addition of CUIs from notes to structured data did not meaningfully improve model performance for predicting clinical deterioration.

**Lay Summary:** Implementing machine learning models to identify clinical deterioration on the wards is associated with improved outcomes. However, these models have high rates of false positives and only use structured electronic health record (EHR) data as predictor variables. Therefore, we aimed to determine if information from clinical notes improves performance and compare methods of combining information from clinical notes with structured data. Model features consisted of variables from structured EHR data (e.g., vital signs, laboratory results) and Concept Unique Identifiers (CUIs) extracted from notes. We parameterized the CUIs in five ways, each of which was combined with the structured data to predict intensive care unit transfer or death in the next 24 hours with a multimodal deep recurrent neural network. The study cohort included 506,076 ward patients across two health systems, 4.9% of whom experienced the outcome. During external validation, the models using both CUIs and structured data performed similarly to the model using only structured data. Of the CUI parameterization approaches, methods that used SapBERT embeddings had the highest discrimination. Our findings show that, while adding medical concept variables from clinical notes could provide additional clinical context for clinicians, they do not enhance model performance compared to structured data alone.

## BACKGROUND AND SIGNIFICANCE

Clinical deterioration, defined as “an acute worsening of a patient’s clinical status that poses a substantial increase to an individual’s short-term risk of death or serious harm,” occurs in up to 5% of hospitalized patients.[1, 2] Early detection of clinical deterioration is essential for minimizing preventable death, as delayed intensive care unit (ICU) transfers and rapid response team activations lead to increased morbidity and mortality.[3–6] Recently, machine learning models developed using large electronic health record (EHR) datasets have been developed to predict clinical deterioration and were shown to have superior performance compared to commonly used tools, such as the Modified Early Warning Score (MEWS) and the National Early Warning Score (NEWS).[1, 7–11] Importantly, implementing machine-learning models for clinical deterioration has been associated with decreased mortality.[12, 13]

Although machine-learning-based early warning scores are more accurate than simpler scores, they still suffer from high false positive rates (FPR), and their input is often restricted to structured EHR variables like vital signs and laboratory results.[1, 11] Consequently, the majority of EHR data, comprised of unstructured text, are not utilized for predicting clinical deterioration. Incorporating information from unstructured text could improve model performance and provide additional clinical context for clinicians by highlighting medical terms that increase deterioration risk. Several methods of combining information from structured data and unstructured clinical notes have been shown to enhance performance in other medical domains.[14–21] However, it is unknown which approach works best when predicting clinical deterioration in hospitalized patients outside the ICU.

## OBJECTIVE

We aim to compare the performance of methods that utilize information from unstructured clinical notes for use in multimodal modeling of clinical deterioration. We also aim to compare these methods to a model that uses only structured data to determine if information for clinical notes improves performance.

## MATERIALS & METHODS

### Patient Population and Data Collection

Adults (age ≥ 18) admitted to the medical-surgical wards at the University of Chicago (UC) from 2016-2022 and the University of Wisconsin-Madison (UW) from 2009-2020 with clinical notes data were eligible for inclusion. Our primary outcome was clinical deterioration, defined as death or direct ward to ICU transfer within 24 hours of each observation, with UC used for model development and UW for external validation.

Patient data were retrieved from the enterprise data warehouses of each university’s health system, which included demographics, vital signs, laboratory values, and clinical notes. All data from UC were de-identified in accordance with the Health Insurance Portability and Accountability Act and transferred to UW for analysis. This study was approved by the Institutional Review Boards of UW (#2019-1258) and UC (#18-0447).

### Structured Features

Fifty-five structured predictor variables were used to develop our models, including demographics, vital signs, laboratory results, and nurse documentation (Supplementary Table 1). For most predictor variables, missing values were imputed by carrying forward in time the last known value, and the remaining missing values were imputed in the development and external validation cohorts using the variable’s median value across all patients in the development cohort. The last known value was only carried forward 24 hours before their missing values were imputed for lactate and blood gas labs. Piecewise-linear encoding (PLE) transformation, a method demonstrated to improve the performance of deep learning models in large, numerical datasets, was used to create a higher-dimensional representation of the data.[22] An additional “hours since admission” input feature was created to capture temporal information.

### Unstructured Input Parameterization

Clinical notes data were pre-processed using the Apache clinical Text Analysis and Knowledge Extraction System (cTAKES), a tool that maps medical terms from the National Library of Medicine’s Unified Medical Language System (UMLS) to Concept Unique Identifiers (CUIs).[23, 24] For example, the term “headache” maps to the CUI “C0018681.” CUIs enable us to focus on the medical terms contained within the notes and create a standard structure to represent notes from distinct health systems. To keep the CUI vocabularies consistent between the UC and the UW data, only CUIs present in the UC development data were kept in the UW data. Furthermore, these CUIs were subset to those appearing in at least five admissions in the UC data. The CUIs were timestamped with the datetimes from their clinical note of origin, then matched with the structured data by pulling forward the most recent 360 unique CUIs that appeared in the clinical notes prior to each structured timestep. 360 was chosen due to computational limitations. Deep learning models cannot operate on CUIs themselves because they are strings. Thus, we investigated various approaches for parameterizing CUIs into model-ready numerical representations (Figure 1).

**Figure 1.**
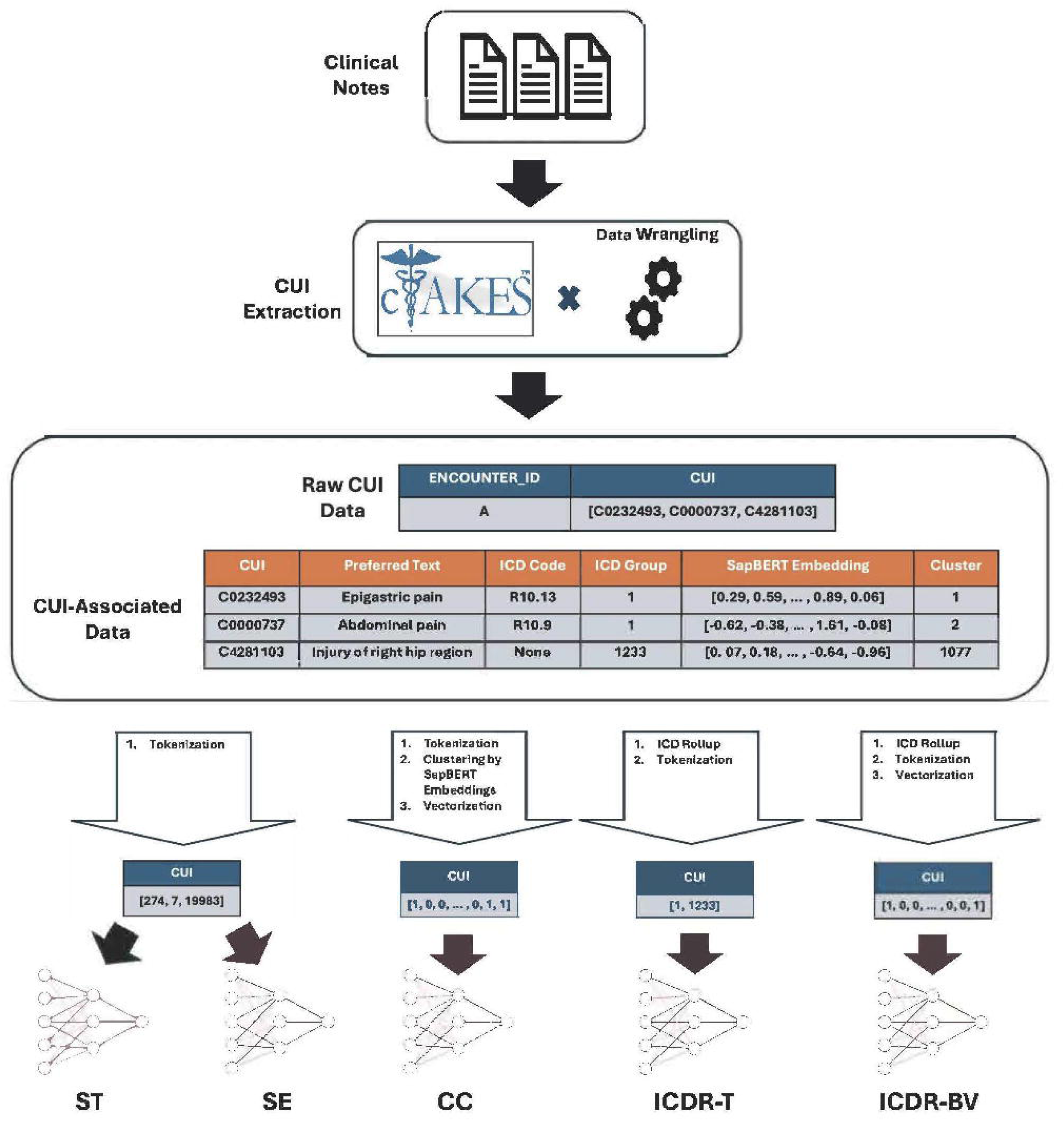
Overview of unstructured data processing for a single timestep. CUIs are extracted from clinical notes by cTAKES and assembled into a usable data structure that maps encounter ID to a list of CUIs associated with the timestep. A table of CUI-associated data is created, independent of patient data, containing preferred text, ICD Code, SapBERT embedding, and cluster information. The CUI-associated data is transformed according to the form required for each model, denoted by the labels beneath each neural network. Note: The data shown are meant to illustrate the processing pipeline and do not have the exact values of patient or CUI-associated data. Abbreviations: cTAKES = Apache clinical Text Analysis and Knowledge Extraction System; CUI = concept unique identifier; ST = standard tokenization; SE = CUIs as SapBERT embedding; CC = CUI clustering using SapBERT embeddings; ICDR-T = ICD rollup using tokenization; ICDR-BV = ICD rollup using binary variables.

#### Standard Tokenization (ST)

The first method of CUI parameterization used a Keras TextVectorization layer to tokenize a list of CUIs into a vector of integers, where there was a unique one-to-one mapping from CUI to integer.

These integers get mapped to dense vectors. In total, 31,418 unique CUIs were shared between the UC and the UW data.

#### ICD Rollup using Tokenization (ICDR-T)

In a second strategy designed to make the CUI inputs more parsimonious, we grouped CUIs capturing similar medical concepts based on the ICD codes associated with them in the UMLS Metathesaurus.[24] Because ICD codes are hierarchical in nature, we used them to “roll up” CUIs whose ICD codes share the first three characters into a unique ICD category. All CUIs without an associated ICD code were assigned to the same category. Mirroring the ST approach, each grouping of CUIs by ICD codes was then mapped to a unique integer. The ICD Rollup groupings were first computed on the UC CUI data and then applied to the UW CUI data. Compared to the ST approach, this process reduced the number of unique tokens from 31,418 to 1,232.

#### ICD Rollup using Binary Variables (ICDR-BV)

Using the tokenized CUI data created in the ICDR-T approach, we converted each vector of tokens to a 1,232-dimensional sparse binary vector, where each index of the sparse vectors represents the presence or absence of a CUI contained by the ICD Rollup grouping corresponding to that index.

#### CUIs as SapBERT Embeddings (SE)

Our next approach to CUI parameterization extended our standard tokenization (ST). We extracted the preferred text strings associated with each CUI (e.g. CUI: “C0000737” → Preferred Text: “Unspecified abdominal pain”) and obtained a 768-dimensional embedding for each string using Hugging Face’s SapBERT model, a transformer model trained on UMLS medical terms.[25, 26] We then generated a mapping from Token → SapBERT Embedding that served as the basis for a pre-trained embedding matrix. When using this embedding approach in our models, the vector of tokens at each timestep is transformed into a matrix of SapBERT embeddings, where there is a one-to-one correspondence between the token value and the row of the embedding matrix that contains that token’s SapBERT embedding (Figure 2).

**Figure 2.**
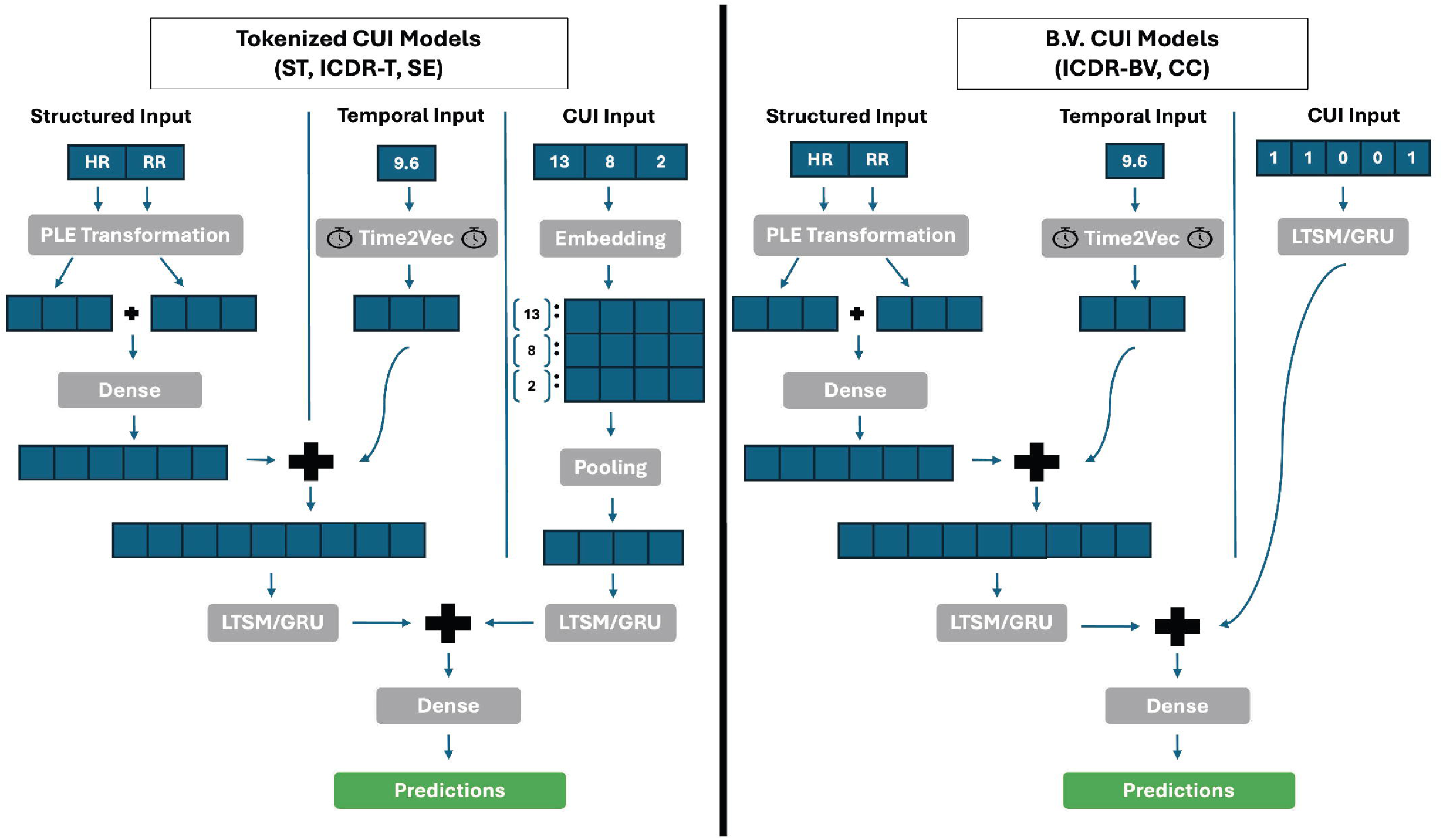
Model architectures for tokenized vs. binary variable (B.V.) CUIs. Each timestep has three inputs: structured, temporal, and unstructured (CUI). The structured and temporal inputs undergo the same transformations in models using tokenized and binary variable CUIs. The structured input is passed through a PLE layer followed by a dense linear layer. The temporal input is vectorized by a Time2Vec layer, before being concatenated with the output of the structural input’s dense layer. The concatenated structured and temporal data are then passed through an LSTM/GRU layer. For models using tokenized CUIs, the list of tokenized CUIs is embedded using either randomly initialized embeddings or SapBERT embeddings, pooled into a single embedding, then passed through an LSTM/GRU layer. For models using binary variable CUIs, the vector of binary CUI variables is directly passed through an LSTM/GRU layer. The LSTM/GRU outputs from the structured + temporal and CUI pipelines are concatenated and then passed through a final dense layer to get the model’s predictions. Dropout layers are excluded for simplicity. Abbreviations: CUI = concept unique identifier; B.V. = binary variables; ST = standard tokenization; SE = CUIs as SapBERT embedding; CC = CUI clustering using SapBERT embeddings; ICDR-T = ICD rollup using tokenization; ICDR-BV = ICD rollup using binary variables; HR = heart rate; RR = respiratory rate; PLE = piecewise linear encoding; LSTM = long short-term memory; GRU = gated recurrent unit.

#### CUI Clustering using SapBERT Embeddings (CC)

Our final approach to CUI parameterization utilized the CUI’s SapBERT embeddings to cluster the CUIs. First, we performed PCA on the embeddings to reduce their dimensionality from 768 to 100. Then, we calculated the pairwise cosine distances between the 100-dimensional embeddings. Lastly, we performed hierarchical clustering with a distance threshold to obtain 1,077 clusters.

With these embedding clusters, we created clusters of tokenized CUIs. These token clusters enabled us to transform the vectors of tokenized CUIs from the ST CUI data into a 1,077-dimensional sparse vector. Like the ICDR-BV approach, each index of the embedding represents the presence or absence of a CUI contained within the cluster associated with that index.

### Model Development

For each CUI parameterization method, a model was developed using an intermediate fusion architecture with long short-term memory (LSTM) or gated recurrent unit (GRU) layers that learned representations of the structured, unstructured (CUI), and temporal inputs separately before combining them to learn their joint interactions. Both the number and type of recurrent layers were hyperparameters. In all models, the structured data were first passed through a fully connected layer, followed by a dropout layer. The temporal data were passed through a Time2Vec layer, a layer for vectorizing time shown to improve LSTM performance. Time2Vec represents time in a neural network by encoding temporal information using a combination of learned linear and sinusoidal functions, capturing both periodic and non-periodic patterns.[27] The structured and temporal data were then concatenated and passed through the LSTM/GRU layer(s).

The CUI data were processed according to their parameterization method. For the ST and ICDR-T approaches, the tokens were transformed into randomly initialized embeddings, which were then passed through a dropout layer. The resulting embeddings were collapsed into a one-dimensional embedding by average or max pooling, passed through a dropout layer, then passed through LSTM/GRU layer(s). A similar process was also applied to the SE approach, with the difference being that the tokens were mapped to their corresponding SapBERT embedding rather than to a randomly initialized one. For the methods parameterizing CUI data as sparse binary vectors (ICDR-BV & CC), the CUI data were passed directly through a dropout layer, then through LSTM/GRU layer(s).

The learned representations of the structured, temporal, and unstructured (CUI) data were fused into a single joint representation. This joint representation was passed through a final fully connected layer with a sigmoid activation function to produce the model’s predictions. A separate, structured data-only model was fit in the same manner but without the additional CUI data.

For hyperparameter optimization, we used the Keras Bayesian Optimization (BO) Tuner with an 80/20 training/validation split on the model development data (UC). We ran twenty trials with early stopping if the validation Area Under the Receiver Operating Characteristic (AUROC) did not improve by ≥0.005 for five consecutive epochs. Batches of size 32 were used for all models, except for the SE model, whose batch size was 16 to not exhaust GPU resources. The external validation data (UW) was not used during model development.

### Model Evaluation

Model discrimination was assessed using the Area Under the Precision-Recall Curve (AUPRC) and the Area Under the Receiver Operating Characteristic (AUROC) as primary (due to the low outcome prevalence) and secondary performance metrics, respectively. AUPRCs with 95% confidence intervals were calculated using bootstrapping with 1000 iterations, where each bootstrapping iteration was done using a sample size equal to 20% of the population size of each subgroup. AUROCs with 95% confidence intervals were calculated using R (version 4.4.0) with the pROC (version 1.18.5) package. Sensitivity, specificity, positive predictive value, and negative predictive value were calculated across a range of cut points.

Subgroup analyses were conducted to assess model performance across sex, race, ethnicity, and age. Differences in patient characteristics were assessed using Chi-squared tests for categorical variables and Mann-Whitney U tests for age.

Initial data cleaning was done using Stata (version 16.1). Preprocessing, descriptive analysis, and model development were done using Python (version 3.9.18), with data analysis and machine learning libraries such as TensorFlow (version 2.12.0), Keras (version 2.12.0), KerasTuner (version 1.3.5), scikit-learn (version 1.2.0), SciPy (version, 1.10.1), and Pandas (version 1.5.3).

## RESULTS

### Cohort Characteristics

The model development cohort (UC) included 284,302 patients, with 14,954 (5.3%) deteriorating during their hospitalization. The external validation cohort (UW) included 248,055 patients, with 11,327 (4.6%) deteriorating during their hospitalization. Across both cohorts (UC and UW), patients who experienced deterioration were more likely to be male (54.7% vs. 48.4% female), older, and have a greater median length of stay than those who did not experience deterioration (Supplementary Table 2). Other demographic variables were statistically significant between those who did and did not experience deterioration, but the magnitudes of these differences were small.

Patient characteristics also varied between sites (Table 1). The model development site (UC) had a higher proportion of females (57.9% vs. 48.4%) and a lower proportion of white patients (39.3% vs. 90.8%). Other differences were statistically significant due to the large sample size but were numerically small.

**Table 1.**
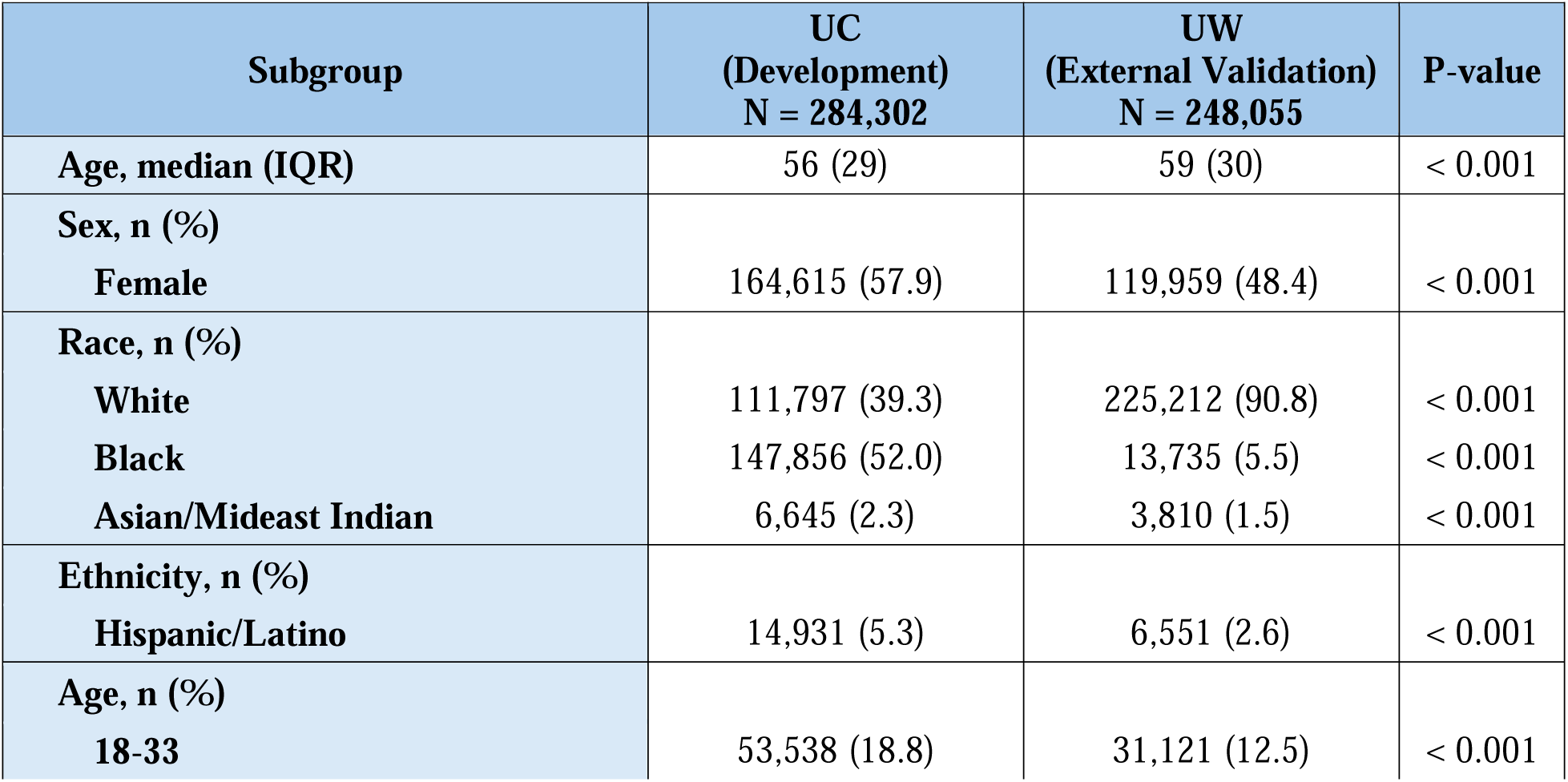
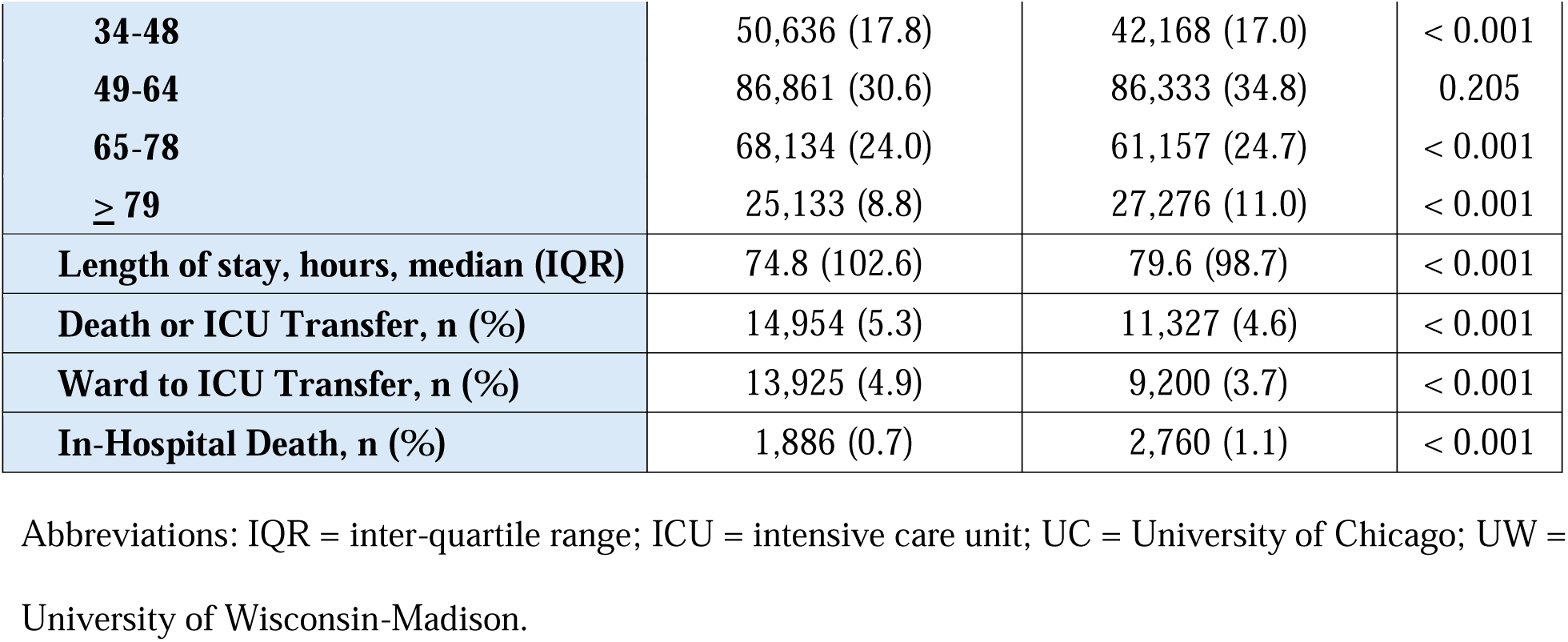
Comparison of patient characteristics between the model development (UC) and external validation (UW) cohorts.

### Model Performance

The AUPRC scores with their 95% CI for predicting clinical deterioration are presented for each model in Supplementary Table 3. The SE model had the highest AUPRC (0.208), followed by the structured-only (0.199) and CC (0.199) models, ICDR-BV (0.194), ICDR-T (0.166), and ST (0.158). The structured-only and CC models achieved the highest AUROC (0.870), followed by ICDR-T (0.867), ICDR-BV (0.866), ST (0.860), and SE (0.859) (Table 2, Figure 3).

**Figure 3.**
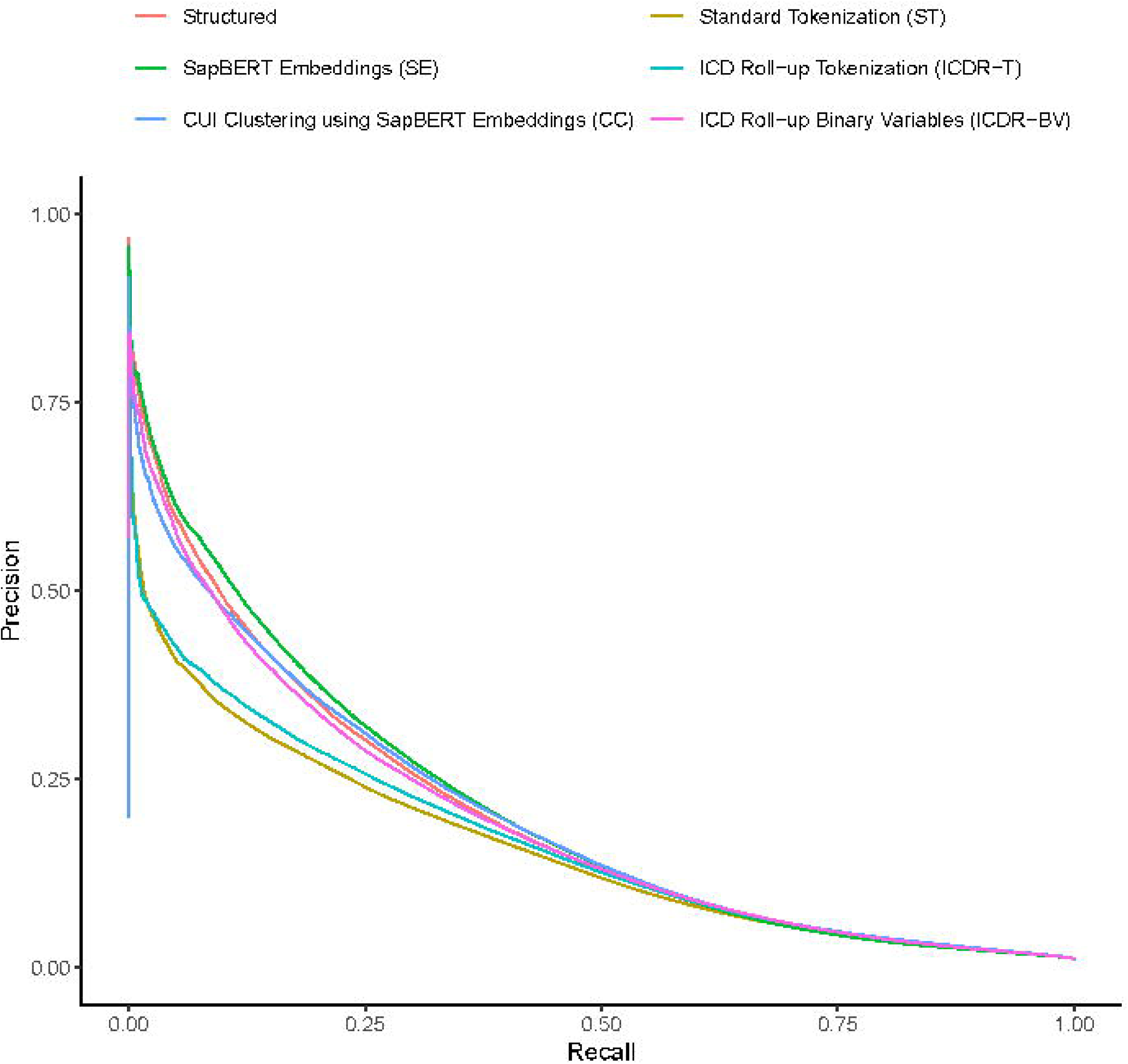
Area under the precision-recall curves (AUPRCs) for the multimodal and structured-only models in the external validation cohort.

**Table 2.** Model AUROCs on external validation cohort (UW) across subgroups.

| Subgroup | Structured<br>AUROC<br>(95% CI) | ST<br>AUROC<br>(95% CI) | ICDR-T<br>AUROC<br>(95% CI) | ICDR-BV<br>AUROC<br>(95% CI) | SE<br>AUROC<br>(95% CI) | CC<br>AUROC<br>(95% CI) |
| --- | --- | --- | --- | --- | --- | --- |
| All | **0.870**<br>(0.869-0.871) | 0.860<br>(0.859-0.861) | 0.867<br>(0.866-0.868) | 0.866<br>(0.865-0.866) | 0.859<br>(0.858-0.859) | **0.870**<br>(0.869-0.871) |
| Sex: Female | 0.861<br>(0.860-0.862) | 0.869<br>(0.868-0.870) | 0.873<br>(0.871-0.874) | 0.873<br>(0.872-0.874) | 0.867<br>(0.866-0.868) | **0.875**<br>(0.874-0.876) |
| Race: White | **0.870**<br>(0.866-0.873) | 0.859<br>(0.858-0.860) | 0.866<br>(0.865-0.867) | 0.865<br>(0.864-0.865) | 0.857<br>(0.856-0.858) | 0.869<br>(0.868-0.870) |
| Race: Black | 0.873<br>(0.867-0.879) | 0.869<br>(0.866-0.872) | 0.870<br>(0.866-0.874) | 0.874<br>(0.871-0.878) | 0.871<br>(0.868-0.875) | **0.881**<br>(0.878-0.884) |
| Race: A/MI | 0.844<br>(0.839-0.850) | 0.866<br>(0.860-0.873) | **0.875**<br>(0.869-0.882) | 0.873<br>(0.867-0.880) | 0.871<br>(0.865-0.878) | 0.872<br>(0.866-0.879) |
| Ethn: Hisp/Lat | **0.862**<br>(0.859-0.865) | 0.849<br>(0.844-0.854) | 0.853<br>(0.847-0.858) | 0.858<br>(0.853-0.863) | 0.852<br>(0.847-0.858) | 0.858<br>(0.853-0.863) |
| Age: 18-33 | **0.871**<br>(0.869-0.873) | 0.867<br>(0.864-0.870) | 0.863<br>(0.860-0.866) | 0.863<br>(0.860-0.866) | 0.854<br>(0.851-0.857) | **0.871**<br>(0.868-0.874) |
| Age: 34-48 | 0.868<br>(0.867-0.869) | 0.873<br>(0.871-0.875) | 0.876<br>(0.875-0.878) | 0.876<br>(0.874-0.878) | 0.870<br>(0.868-0.872) | **0.882**<br>(0.881-0.884) |
| Age: 49-64 | 0.847<br>(0.846-0.849) | 0.862<br>(0.861-0.864) | 0.872<br>(0.871-0.874) | 0.870<br>(0.869-0.871) | 0.864<br>(0.863-0.866) | **0.875**<br>(0.874-0.876) |
| Age: 65-78 | 0.846<br>(0.844-0.848) | 0.846<br>(0.844-0.847) | 0.854<br>(0.852-0.855) | 0.853<br>(0.852-0.855) | 0.843<br>(0.841-0.844) | **0.857**<br>(0.856-0.858) |
| Age: ≥ 79 | **0.862**<br>(0.861-0.863) | 0.845<br>(0.843-0.847) | 0.851<br>(0.849-0.853) | 0.850<br>(0.848-0.852) | 0.844<br>(0.842-0.846) | 0.853<br>(0.851-0.855) |

The best score for each subgroup is bold. The worst score for each subgroup is italicized.
Abbreviations: AUROC = area under the receiver operating characteristic; CI = confidence interval; A/MI = Asian/Mideast Indian; Ethn = ethnicity; Hisp/Lat = Hispanic/Latino; ST = standard tokenization; SE =
CUIs as SapBERT embedding; CC = CUI clustering using SapBERT embeddings; ICDR-T = ICD rollup using tokenization; ICDR-BV = ICD rollup using binary variables.

Some variation of model performance existed across subgroups. Across all models, the AUPRC scores for Asian/Mideast-Indian (A/MI) patients were the greatest on average (0.224), while they were the lowest on average for patients between the ages of 18-30 (0.156). AUROC scores were more tightly clustered than AUPRC scores across subgroups. Still, the average AUROC score across models was greatest for patients in the age range of 34-48 (0.875), while it was the lowest for patients of age >= 79 (0.849). The SE model had the highest performance across all subgroups for AUPRC, while the best-performing model differed across subgroups for AUROC.

Supplementary Table 4 depicts the sensitivity, specificity, positive, and negative predictive values for models assessed along a range of probability cutoffs corresponding to the highest-risk 15%, 10%, 5%, and 1% of observations for clinical deterioration. At the 5% cutoff, the CC model achieved the greatest positive predictive value (12.53%) and sensitivity (52.15%). At the 15% cutoff, the ICDR-T, CC, and ICDR-BV models tied for the highest positive predictive value at 5.67%, while their sensitivities were 70.95%, 70.92%, and 70.86%, respectively. The average positive predictive values across models at the 5% and 15% cutoffs were 12.30% and 5.63%, respectively, while the average sensitivities were 51.23% and 70.27%.

## DISCUSSION

In this study, we compared five methods of CUI parameterization for use in multimodal deep learning models as well as a structured-only model to predict clinical deterioration in non-ICU inpatients. The performance of all models was strong, and discrimination was similar overall, especially with the AUROC metric. We found no meaningful differences between models that utilized tokenized + embedded CUI data, like the SE, ICDR-T, and ST models, versus those that included CUI data as a vector of binary variables, like ICDR-BV and CC models. However, our method utilizing SapBERT embeddings to represent CUIs (SE) achieved the highest AUPRC overall and across all subgroups. However, this performance was only slightly better than the structured data-only model. Our results suggest that the addition of medical terms as CUIs in multimodal models does not meaningfully improve performance beyond models using structured data alone.

To our knowledge, this is the first paper to investigate multimodal integration of information from structured and unstructured data in deep learning models to predict clinical deterioration in ward patients. A recent systematic review by Van Der Vegt et al. examined the current use of artificial intelligence for predicting clinical deterioration. They found that while many groups have developed promising machine learning models using methods such as logistic regression, XGBoost, and random forests, few have developed deep learning models.[28] Deep learning models have, on average, outperformed non-deep learning models.[29–31] Also, groups have developed deep learning models for predicting deterioration using ICU patients, patients from the emergency department, or patients with COVID-19.[10, 21, 32–35] However, few have developed models using ward patients.[11, 29] Fewer still have utilized multiple input modalities, none of which focused on ward deterioration.[36] Because predicting clinical deterioration early and accurately leads to improved outcomes, efforts to increase model accuracy like ours could lead to enhanced detection and decrease false alarms. Furthermore, the incorporation of information from clinical notes could provide additional context for clinicians related to factors that increase a patient’s risk of the event.

When including CUIs in multimodal models, our results suggest a benefit in restricting the number of CUI-related variables in the input space. The CC, ICDR-T, and ICDR-BV models restrict the number of CUI-related variables in their unstructured input from 31,418 to 1,077, 1,232, and 1,232, respectively, while the ST and SE models use the full 31,418. The models restricting the CUI-related variable space achieved higher AUROCs, positive predictive values, and sensitivities at the 15% cutoff relative to SE and ST models. However, this pattern does not hold as strongly for AUPRC scores. The ST model, which does not restrict the number of CUI-related variables, achieved a relatively low AUPRC, as it did for AUROC, but the SE model, which achieved the lowest AUROC, achieved the highest AUPRC score. When considering all performance metrics together, the CC model may have a slight edge above the other CUI models, as it achieved the highest AUROC, positive predictive values, and sensitivities at the 5% cutoff, the highest positive predictive value (tie) at the 15% cutoff, and the second highest AUPRC. However, all differences in performance metrics were small. Importantly, the optimal model for clinical implementation would be based on the cutoff threshold picked that would prompt specific clinical actions or alerts needed for clinical practice. We also found that the structured-only model performed just as well and often better than most of the models that used CUIs. Thus, the higher complexity of implementing and interpreting the CUI models must be weighed against the additional clinical context information that could be provided by these models (e.g., using explainable artificial intelligence techniques) when considering whether to use these models in practice.

Our methods also demonstrate the ability to make effective and fair predictions in ward patients. Existing papers developing deep learning models in ward patients did not provide analyses on the performance of their models across patient subgroups, which is critical given the diverse nature of ward patients.[11, 29, 36] Our models obtained strong performance on patients with diverse demographics coming from variable hospital settings. AUROCs were slightly higher on average for patients aged 34-48 when compared to other subgroups, and slightly worse for patients near the tail ends of the age distribution (18-33 and ≥79). Furthermore, the model discrimination was superior for black patients when compared to other racial subgroups. This might be explained by the high proportion (52.0%) of black patients in the development (UC) cohort. Additionally, the AUPRC scores for patients aged 18-33 were uniformly the lowest across all models. A likely cause for this may be that the proportion (7.3%) of these patients was lower than any other subgroup.

Our study has several strengths. First, incorporating multimodal data into deep learning models is rare within the ward deterioration literature, as most groups who have developed deep learning models have used exclusively structured EHR data.[10, 28–30, 33, 34, 37–39] Our study provides a diverse set of methods to inspire further exploration of multimodal modeling of clinical deterioration. Second, each CUI parameterization method and its associated model’s performance were externally validated in a separate health system. Large discrepancies existed between the development and external validation cohorts, and note-taking practices can differ greatly between hospital systems, which could increase variability in the CUIs for similar patients.[40–42] Despite these challenges, all models exhibited strong performance, suggesting the robustness and generalizability of our methods to diverse settings.

Our study is limited in that we did not explore all possible CUI parameterization methods, and we only focused on predicting a single outcome. Also, this study did not use the raw text that constitutes clinical notes. Thus, we cannot draw strong conclusions about the overall utility of clinical notes in clinical prediction models. Finally, this was a retrospective study, and the usefulness of a clinical prediction model needs to be tested in a prospective clinical trial, which is outside the scope of this work.

## CONCLUSION

In conclusion, we demonstrated the feasibility of combining structured and unstructured EHR data in deep learning models to predict clinical deterioration. While the best-performing model varied by metric, those whose method of CUI parameterization utilized SapBERT embeddings may have an edge over others, as the SE model performed best with respect to our primary metric (AUPRC), while the CC model seemed to perform best when considering all metrics in aggregate, achieving the highest AUROC, second highest AUPRC, and highest positive predictive value at multiple cutoffs. However, the structured-only model performed similarly to the models that included CUIs, suggesting that the marginal gains in performance may not outweigh the increased complexity and difficulties in interpreting CUI models. This work adds to the field of predicting clinical deterioration, which has historically only focused on structured data.

## Data Availability

The data utilized in this article cannot be shared publicly because of legal and regulatory restrictions. These data were obtained from four hospital systems after our research protocol was reviewed by IRBs from each hospital, and our data use agreements do not permit sharing due to the granular nature of the data.

## Acknowledgments

None

## Author Contributions

Matthew M. Churpek, Majid Afshar, and Anoop Mayampurath conceptualized the study. Matthew M. Churpek, Majid Afshar, and Dana P. Edelson managed data collection and secured IRB approvals. Kyle A. Carey, Jennifer Martin, and Charles A. Kotula carried out the data preprocessing. Charles A. Kotula, Jennifer Martin, Matthew M. Churpek, Majid Afshar, Anoop Mayampurath, and Dmitriy Dligach are responsible for data analysis and methodology. Charles A. Kotula prepared the original draft, and all authors reviewed and edited the manuscript.

## Institutional Review Boards

Ethical approval of this work was given by the University of Wisconsin-Madison Minimal Risk Research IRB (#2019-1258) and the University of Chicago Biological Sciences Division IRB (#18-0447).

## Data Sharing Statement

The data used in this study were acquired from two hospital systems following approval from the IRBs. The data use agreements prohibit sharing data due to regulatory and legal constraints, and therefore, the data cannot be shared publicly.

