## Supplementary material for "Comparison of Multimodal Deep Learning Approaches for Predicting Clinical Deterioration in Ward Patients": ONLINE DATA SUPPLEMENT

**Supplementary Table 1.** Predictor variables used in the models.

| **Demographics** | Age | Continuous |
| --- | --- | --- |
| **Vital signs** | Temperature (C°), Heart Rate, Respiratory Rate, Systolic Blood Pressure (SBP), Diastolic Blood Pressure (DBP), O2 Saturation, Fraction of Inspired Oxygen (FiO2), AVPU, Disorientation (yes/no) | Continuous |
| **Laboratory values** | Basic Metabolic Panel [BMP]: Sodium, Chloride, Potassium, Bicarbonate (CO2), Anion Gap, Glucose, Calcium, Blood Urea Nitrogen (BUN), Serum Creatinine (SCr), Phosphate | Continuous |
|  | Liver Function Test [LFT]: Total Protein, Albumin, Total Bilirubin, Aspartate Aminotransferase (AST/SGOT), Alkaline Phosphatase |  |
|  | Complete Blood Count [CBC]: White Blood Cells (WBC), Hemoglobin, Platelet Count, Bands, Eosinophils, Lymphocytes, Monocytes, Neutrophils |  |
|  | Blood Gas test: Arterial pH, Venous pH, Arterial Partial Pressure of Oxygen, Arterial Partial Pressure of Carbon Dioxide, Venous Partial Pressure of Carbon Dioxide |  |
|  | Other labs: Lactate, Magnesium, Lipase, International Normalized Ratio (INR), Mean Corpuscular Volume (MCV), Partial Thromboplastin Time (PTT), and Red Cell Distribution Width (RDW) |  |
| **Nurse Documentation** | Braden Scale (Activity, Friction and Shear, Mobility, Moisture, Nutrition, Sensory Perception, Total Score), Body Mass Index | Continuous |
| **Length of stay** | Hours since admission until the current time point | Continuous |
| **Time of Day** | Hours since midnight of the current day | Continuous |
| **Urinary Output** | Sum of Urine Output over the last 24 hours | Continuous |

**Supplementary Table 2.** Comparison of patient characteristics between those with and without clinical deterioration in the study cohort.

| **Characteristic** | **Deterioration (n= 26,281)** | **No Deterioration (n= 506,076)** | **P-value** |
| --- | --- | --- | --- |
| **Age, median (IQR)** | 63 (21) | 57 (27) | < 0.001 |
| **Female, n (%)** | 11,910 (45.3) | 272,664 (53.9) | < 0.001 |
| **Race, n (%)** |  |  |  |
| **White** | 16,320 (62.1) | 320,689 (63.4) | < 0.001 |
| **Black** | 8,231 (31.3) | 153,360 (30.3) | < 0.001 |
| **Asian/Mideast Indian** | 500 (1.9) | 9,955 (2.0) | < 0.001 |
| **Hispanic/Latino ethnicity, n (%)** | 1,010 (3.8) | 20,472 (4.0) | < 0.001 |
| **Age, n (%)** |  |  |  |
| **18-33** | 1,924 (7.3) | 82,735 (16.3) | < 0.001 |
| **34-48** | 3,238 (12.3) | 89,566 (17.7) | < 0.001 |
| **49-64** | 9,194 (35.0) | 164,000 (32.4) | < 0.001 |
| **65-78** | 8,298 (31.6) | 120,993 (23.9) | < 0.001 |
| **≥ 79** | 3,627 (13.8) | 48,782 (9.6) | < 0.001 |
| **Length of stay, hours, median (IQR)** | 217.5 (294.6) | 74.2 (93.7) | < 0.001 |

Abbreviations: IQR = inter-quartile range.

**Supplementary Table 3.** Model AUPRCs on External Validation Cohort (UW) Across Subgroups

| **Subgroup** | **Structured  AUPRC  (95% CI)** | **ST  AUPRC  (95% CI)** | **ICDR-T  AUPRC  (95% CI)** | **ICDR-BV  AUPRC  (95% CI)** | **SE   AUPRC  (95% CI)** | **CC  AUPRC  (95% CI)** |
| --- | --- | --- | --- | --- | --- | --- |
| **All** | 0.199 (0.196-0.203) | *0.158 (0.155-0.161)* | 0.166 (0.163-0.169) | 0.194 (0.191-0.198) | **0.208 (0.204-0.211)** | 0.199 (0.195-0.202) |
| **Sex: Female** | 0.197 (0.191-0.202) | *0.159 (0.155-0.164)* | 0.165 (0.160-0.170) | 0.193 (0.188-0.199) | **0.205 (0.200-0.211)** | 0.195 (0.189-0.200) |
| **Race: White** | 0.197 (0.193-0.200) | *0.156 (0.153-0.159)* | 0.165 (0.162-0.169) | 0.192 (0.188-0.195) | **0.205 (0.201-0.209)** | 0.197 (0.193-0.200) |
| **Race: Black** | 0.217 (0.200-0.234) | 0.183 (0.169-0.200) | *0.179 (0.165-0.195)* | 0.209 (0.193-0.226) | **0.225 (0.209-0.242)** | 0.215 (0.199-0.234) |
| **Race: A/MI** | 0.237 (0.201-0.270) | *0.187 (0.160-0.219)* | 0.196 (0.168-0.228) | 0.239 (0.208-0.273) | **0.244 (0.210-0.277)** | 0.252 (0.218-0.285) |
| **Ethn: Hisp/Lat** | 0.228 (0.204-0.255) | *0.174 (0.154-0.197)* | 0.197 (0.176-0.222) | 0.223 (0.199-0.247) | **0.244 (0.220-0.272)** | 0.227 (0.205-0.254) |
| **Age: 18-33** | 0.173 (0.160-0.187) | *0.130 (0.120-0.141)* | 0.146 (0.135-0.158) | 0.166 (0.153-0.179) | **0.174 (0.160-0.188)** | 0.165 (0.153-0.179) |
| **Age: 34-48** | 0.203 (0.193-0.213) | *0.163 (0.154-0.172)* | *0.163 (0.154-0.171)* | 0.196 (0.186-0.206) | **0.214 (0.203-0.225)** | 0.201 (0.190-0.211) |
| **Age: 49-64** | 0.211 (0.205-0.217) | *0.161 (0.156-0.165)* | 0.169 (0.164-0.174) | 0.203 (0.198-0.209) | **0.223 (0.216-0.229)** | 0.215 (0.209-0.221) |
| **Age: 65-78** | 0.191 (0.185-0.196) | *0.153 (0.148-0.157)* | 0.165 (0.160-0.170) | 0.189 (0.184-0.195) | **0.199 (0.193-0.204)** | 0.189 (0.183-195) |
| **Age: >= 79** | 0.196 (0.186-0.205) | *0.181 (0.172-0.190)* | 0.186 (0.177-0.195) | 0.192 (0.183-0.201) | **0.200 (0.191-0.210)** | 0.195 (0.186-0.204) |

The best score for each subgroup is bold. The worst score for each subgroup is italicized.

Abbreviations: AUPRC = area under the precision-recall curve; CI = confidence interval; A/MI = Asian/Mideast Indian; Ethn = ethnicity; Hisp/Lat = Hispanic/Latino; ST = standard tokenization; SE = CUIs as SapBERT embedding; CC = CUI clustering using SapBERT embeddings; ICDR-T = ICD rollup using tokenization; ICDR-BV = ICD rollup using binary variables.

**Supplementary Table 4.** Sensitivity, specificity, positive predictive value, and negative predictive value for all models in the validation cohort across probability cutoffs of 15%, 10%, 5%, and 1% of observations predicted to have a positive outcome. Rows for each cutoff are presented in decreasing order of sensitivity.

| **Cutoff = 15%** | | | | | | |
| --- | --- | --- | --- | --- | --- | --- |
| **Model** | **Pred. Prob. Cutoff** | **Obs. Above Cutoff (%)** | **SENS** | **SPEC** | **PPV** | **NPV** |
| ICDR-T | 0.018 | 3,589,339 (15.0) | 70.95 | 85.69 | 5.67 | 99.59 |
| CC | 0.009 | 3,586,574 (15.0) | 70.92 | 85.70 | 5.67 | 99.59 |
| ICDR-BV | 0.015 | 3,586,305 (15.0) | 70.86 | 85.70 | 5.67 | 99.59 |
| Structured | 0.020 | 3,583,379 (15.0) | 69.91 | 85.70 | 5.60 | 99.58 |
| ST | 0.019 | 3,854,185 (15.0) | 69.52 | 85.70 | 5.57 | 99.57 |
| SE | 0.020 | 3,582,715 (15.0) | 69.48 | 85.70 | 5.57 | 99.57 |
| **Cutoff = 10%** | | | | | | |
| **Model** | **Pred. Prob. Cutoff** | **Obs. Above Cutoff (%)** | **SENS** | **SPEC** | **PPV** | **NPV** |
| CC | 0.014 | 2,391,102 (10.0) | 63.79 | 90.67 | 7.66 | 99.52 |
| ICDR-BV | 0.023 | 2,391,717 (10.0) | 63.76 | 90.67 | 7.65 | 99.52 |
| ICDR-T | 0.028 | 2,393,387 (10.0) | 63.47 | 90.65 | 7.61 | 99.51 |
| SE | 0.029 | 2,392,567 (10.0) | 62.91 | 90.65 | 7.55 | 99.51 |
| Structured | 0.031 | 2,391,071 (10.0) | 62.66 | 90.65 | 7.52 | 99.50 |
| ST | 0.029 | 2,390,283 (10.0) | 62.21 | 90.65 | 7.47 | 99.50 |
| **Cutoff = 5%** | | | | | | |
| **Model** | **Pred. Prob. Cutoff** | **Obs. Above Cutoff (%)** | **SENS** | **SPEC** | **PPV** | **NPV** |
| CC | 0.033 | 1,194,691 (5.0) | 52.15 | 95.58 | 12.53 | 99.40 |
| SE | 0.057 | 1,196,661 (5.0) | 51.86 | 95.57 | 12.44 | 99.39 |
| ICDR-BV | 0.047 | 1,195,276 (5.0) | 51.56 | 95.57 | 12.38 | 99.39 |
| ICDR-T | 0.058 | 1,196,365 (5.0) | 51.01 | 95.56 | 12.24 | 99.34 |
| Structured | 0.063 | 1,196,168 (5.0) | 50.88 | 95.56 | 12.21 | 99.38 |
| ST | 0.061 | 1,196,371 (5.0) | 49.90 | 95.55 | 11.97 | 99.37 |
| **Cutoff = 1%** | | | | | | |
| **Model** | **Pred. Prob. Cutoff** | **Obs. Above Cutoff (%)** | **SENS** | **SPEC** | **PPV** | **NPV** |
| SE | 0.25 | 239,395 (1.0) | 26.01 | 99.30 | 31.18 | 99.10 |
| CC | 0.19 | 239,287 (1.0) | 25.55 | 99.30 | 30.63 | 99.10 |
| Structured | 0.29 | 239,478 (1.0) | 25.28 | 99.29 | 30.30 | 99.10 |
| ICDR-BV | 0.21 | 239,362 (1.0) | 24.48 | 99.29 | 29.34 | 99.09 |
| ICDR-T | 0.30 | 239,391 (1.0) | 22.70 | 99.26 | 27.22 | 99.06 |
| ST | 0.33 | 239,409 (1.0) | 21.77 | 99.25 | 26.10 | 99.05 |

Abbreviations: ICDR-T = ICD rollup using tokenization; CC = CUI clustering using SapBERT embeddings; ICDR-BV = ICD rollup using binary variables; ST = standard tokenization; SE = CUIs as SapBERT embedding; Pred = predicted; Prob = probability; SENS = sensitivity; SPEC = specificity; PPV = positive predictive value; NPV = negative predictive value.
